# Impairment and characteristics of postural control sub-components in people with COPD: a scoping review

**DOI:** 10.1101/2022.04.13.22273798

**Authors:** Romain Pichon, Olivier Van Hove, Mathieu Ménard, Diane Hearing, Armel Crétual

**Author notes:** Corresponding author: Romain Pichon, Institut de Formation en Pédicurie-Podologie, Ergothérapie et Kinésithérapie (IFPEK), 12 rue Jean Louis Bertrand, 35000 Rennes, France.

## Abstract

**Purpose:** Impairment of postural control is a common extra-respiratory manifestation in people with COPD. However, the precise characteristics of this alteration are not clearly known. The “Systems Framework for Postural Control” which define postural control sub-components, is a relevant tool to explore this field. The main aim of this review was to identify which postural control sub-components are impaired in patients with COPD and to summarize characteristics for each sub-component. A secondary aim was to precise the relation between postural control and activities of daily living.

**Materials and methods:** A scoping review was conducted, according to the JBI methodology. Pubmed, Cochrane Library, Scielo, Google Scholar, Open Grey and HAL were searched from inception to March 2021. The search was performed in English and French.

**Results:** Seventy-seven articles were included. There was evidence of a potential impairment for most of the postural control sub-components. Characteristics of every sub-component alteration were heterogeneous. Even if the relation was poorly studied, reduced postural control seemed to be associate with difficulties in activities of daily living.

**Conclusion:** Patients with COPD could experiment impairment among a wide range of postural control sub-components. Future research must clarify if a common pattern of modification exits for this alteration.

**Implications for rehabilitation:**

- Impairment of postural control is a common extra-respiratory manifestation in patients with COPD and so clinicians must include it in their clinical reasoning
- Numerous postural control sub-components could be altered in patients with COPD, suggesting that postural control assessment must be holistic.
- This scoping review shows that characteristics of postural control impairment are varied and that there may be no common pattern at the COPD population level.

## Introduction

According to the World Health Organization [1], Chronic Obstructive Pulmonary Disease (COPD) is the third worldwide leading cause of mortality in 2019. COPD induces important socio-economic problems and is considered as an international major concern [2,3]. COPD is a common, preventable and treatable pathology that is characterized by persistent respiratory symptoms and airflow limitation [4]. The main respiratory symptoms are dyspnea, cough and sputum hypersecretion [4]. The primary cause is exposure to tobacco smoke and other risk factors include exposure to various inhaled toxics [3,4]. Hence, it is well established that COPD has an important impact on people’s activities and quality of life [5].

Beyond respiratory symptoms, comorbidities are extremely frequent during the evolution of the disease [6]. Presence of comorbidities increase the difficulties in activities of daily living and the impact on the quality of life of COPD patients [7]. Among these comorbidities, the impairment of postural control is one important extra-respiratory manifestation for these patients that could impact their daily lives [8].

The postural control is a complex system, which allows to manage the body’s position in the space for the dual purposes of stability and orientation [9]. Three systematic reviews highlighted the impairment of postural control of COPD compared to non-pathological subjects [10,11,8]. However, the characteristics and the mechanisms of the postural control alteration are still unknown and the impact on daily living activities is poorly known.

Contemporary theories consider the postural control as a global organisation that integrate various inputs and tend to adapt the body as a mechanical system in interaction with the nervous system in a continuously changing environment [9]. This is supported by numerous studies, showing that impairments of these systems can alter the postural control [12]. To evaluate the postural control and better understand the whole implicated system, Horak [13] proposed the *Systems Framework for Postural Control* and identifies different interconnected sub-components involved, that are presented in *table1*.

**Table 1.** Postural control sub-components definitions.

| **Domains in Systems Framework for Postural Control [13]** | **Adaptation by Sibley et al. [16]** | **Operational definition by Sibley et al.[16]** |
| --- | --- | --- |
| 1. Biomechanical constraints | 1. Static stability | Ability to maintain position of the center of mass in unsupported stance when the base of support does not change |
|  | 2. Underlying motor systems | E.g. strength and coordination |
|  | 3. Functional stability limits | Ability to move the center of mass as far as possible in the anteroposterior or mediolateral directions within the base of support |
| 2. Orientation in space | 4. Verticality | Ability to orient appropriately with respect to gravity (e.g. evaluation of lean) |
| 3. Movement strategies | 5. Reactive postural control | Ability to recover stability after an external perturbation to bring the center of mass within the base of support through corrective movements (e.g. ankle, hip and stepping strategies) |
|  | 6. Anticipatory postural control | Ability to shift the center of mass before a discrete voluntary movement (e.g. stepping-lifting leg, arm raise, head turn) |
| 4. Control of dynamics | 7. Dynamic stability | Ability to exert ongoing control of center of mass when the base of support is changing (e.g. during gait and postural transitions) |
| 5. Sensory strategies | 8. Sensory integration | Ability to reweight sensory information (vision, vestibular, somatosensory) when input is altered |
| 6. Cognitive processing | 9. Cognitive influences | Ability to maintain stability while responding to commands during the task or attend to additional tasks (e.g. dual-tasking) |

This framework has been used to develop and classify postural control assessment tools [12,14– 17]. To identify impairments in postural control sub-components in patients with COPD and clarify the role of these alterations in daily living activities for COPD patients, this work proposes a scoping review framed on this model. The main aim was to identify and characterize the alterations of the postural control sub-components in patients with COPD. A second aim proposed to explore the association between the postural control and the difficulties encountered during activities of daily living by patients with COPD.

## Materials and methods

This scoping review was conducted in accordance with the Johanna Briggs Institute (JBI) methodology for scoping review [18]. The objectives, inclusion criteria and methods for this scoping review were specified in advance and documented in an *a priori* protocol, registered on OSF.io the 03-02-2021 [19].

### Inclusion criteria

#### Participants

This review considered studies that include COPD patients as defined by the *Global Initiative for Obstructive Lung Disease* (GOLD) [4]. All stages of the disease were considered. Patients must be over 18 years old. This review excluded studies involving other chronic respiratory lung diseases (eg. Asthma, interstitial lung diseases or bronchiectasis).

#### Concept

The concept of postural control (through the “Systems Framework for Postural Control” proposed by Horak [13]) and his sub-components were explored in this review. This model was used and adapted by Sibley et al. [16] who established an operational definition of the model. The adaptation includes 9 sub-components which are presented and defined in the *table1*.: 1. Static stability, 2. Underlying motor systems, 3. Functional stability limits, 4. Verticality, 5. Reactive postural control, 6. Anticipatory postural control, 7. Dynamic stability, 8. Sensory integration, 9. Cognitive influences.

This review included studies that focus on postural control, or at least one of its sub-components but in relation with global postural control strategies or performance in static or dynamic conditions. This work considered as postural control assessment all functional tools described by Sibley et al. [16] and laboratory assessments (kinetic or kinematic analyses in static and dynamic conditions) [20]. Subjective assessments of postural control were not included in the review. Sources analysing one sub-component in isolation, with no direct link to postural control are excluded from the review.

#### Context

Postural control is now identified as an important concern for the assessment and the management of COPD patients all over the world and in every care setting [11]. This review focused on available evidence on postural control with no geographical restriction. We included studies in the widest possible range of settings (laboratory assessment, inpatient or outpatient studies, pulmonary rehabilitation settings) and population (community-dwelling, real life…).

#### Types of studies

This scoping review considered observational studies (descriptive studies, cohort studies, cross-sectional studies, case-control studies), experimental studies (randomized controlled trials, non-randomized controlled trials, and quasi-experimental designs such as before-after studies) and systematic reviews. Conference abstracts and grey literature were considered for inclusion, as they represent an important source of unpublished evidence that can have an impact on review’s results and conclusions [21,22].

#### Search Strategy

The search strategy follows the three-steps methodology recommended by the JBI. Firstly, an initial search was carried out on PubMed and Cochrane Library, to identify articles related to our research question. The text words included in titles and abstracts and the different keywords were used to develop a complete search strategy. Secondly, the full-search strategy was developed including information from the *first step* and completed by specific keywords related to each postural control sub-components. The search was conducted on PubMed, Google Scholar, Scielo and Cochrane Library. The full search strategy for PubMed is presented in *Appendix I*. Search strategy was adapted as necessary for each database. For the search on Google Scholar, the 500 first results were screened. Finally, references list of identified articles were searched for additional sources.

A search for grey literature was conducted on OpenGrey and the multidisciplinary open archive HAL. If relevant, reviewers (RP and OVH) contacted authors for further information.

Studies published in English and French were considered for inclusion. The databases were searched from inception to March 3, 2021.

#### Source of evidence selection

All identified citations were uploaded into *Rayyan* application. Two researchers (RP and OVH) independently reviewed titles, abstracts, and keywords from identified citations and applied inclusion and exclusion criteria. In the case of incomplete information, full text was studied. Then, the same two researchers examined independently the full texts applying inclusion and exclusion criteria. In the case of disagreement, an argumentative discussion was implemented between the two reviewers and a third member of the team was consulted if the discussion does not permit a decision. Reasons for exclusion of sources that do not meet inclusion criteria were recorded and presented in the final review. The complete results of the search were reported in the final report and presented in a Preferred Reporting Items for Systematic Reviews and Meta-Analysis extension for Scoping Reviews (PRISMA-ScR) flow diagram [23].

#### Data extraction

Relevant data were extracted from papers included in this scoping review using a charting table developed in connection to the questions and the objectives of the review. A pilot testing of the charting table was conducted. As planned in the *a priori* protocol, the charting table was modified after the pilot testing by the reviewers. The final version of the charting table is available in the *appendix II*.

## Results

### Study inclusion

Our searches returned 10313 records. Following the exclusion of 2902 duplicated records, 7415 records were screened by title and abstracts. 7218 records that did not meet the inclusion criteria were then excluded and 197 records were thought for full text retrieval. For 31 sources, full texts were not available and full text examination was performed on 166 studies. 89 sources did not meet the inclusion criteria of this scoping review, leaving a final 77 sources eligible for the review. The screening process was reported in a PRISMA-Scoping Review study flow diagram (*figure*.*1*).

**Figure 1.**
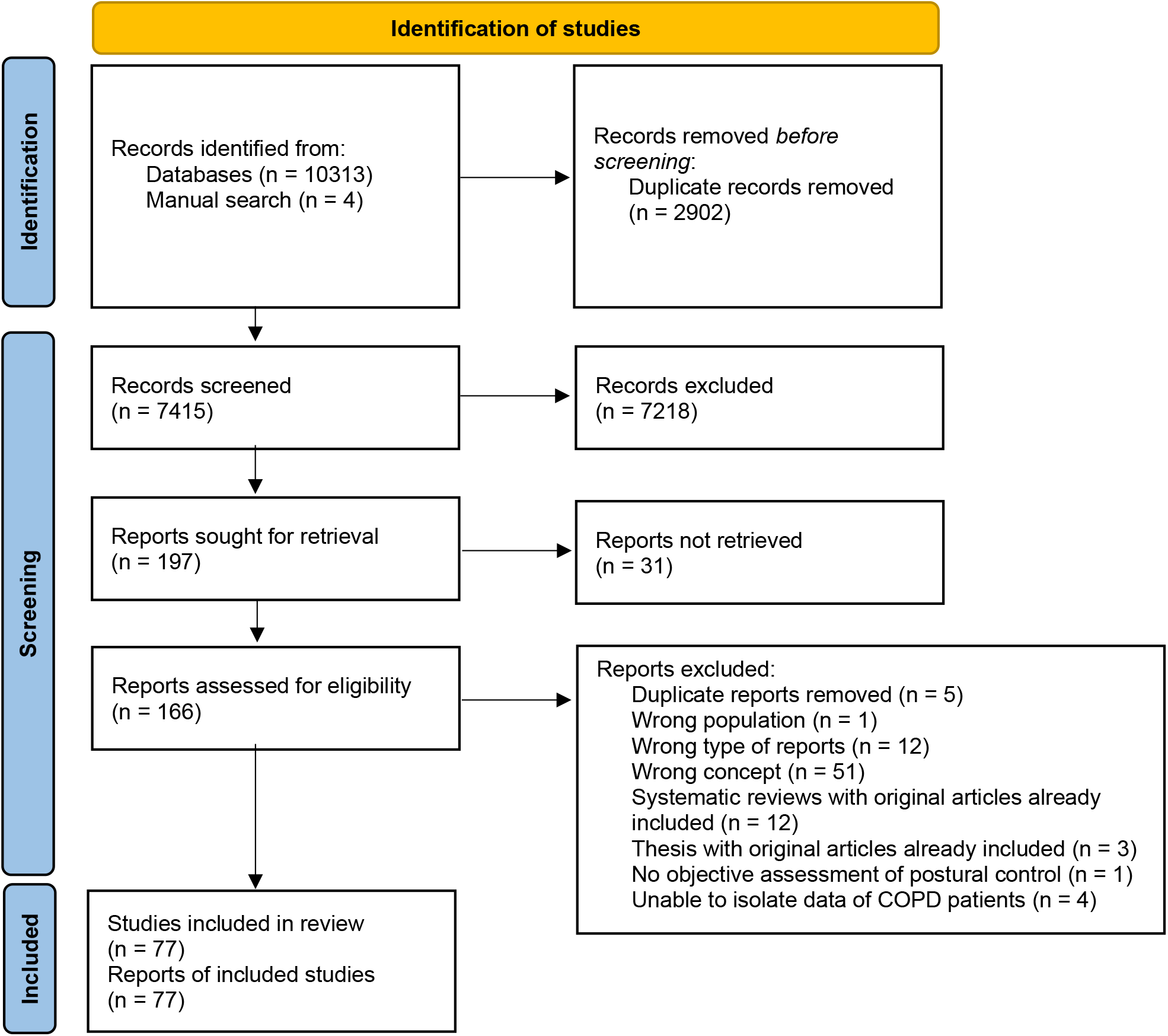
PRISMA-Scoping Review flow diagram. flow diagram representing the successive steps of studies identification, screening, and inclusion for the scoping review. After the full process, 77 studies were included in the review. Long description: flow diagram representing the successive steps of studies identification, screening, and inclusion for the scoping review. The first step was the initial search that identified a total of 10317 sources. After deduplication, the second step consisted in the screening by the reviewers of 7415 records. The application of eligibility criteria resulted in 77 studies that were included in the scoping review.

### Characteristics of included studies

#### Year of publication

The earliest study included in this review was published in 2004. There is a trend toward an increase of the number of publications from 2012 (3 studies) until a peak in 2020 (15 studies) (*figure*.*2*).

**Figure 2.**
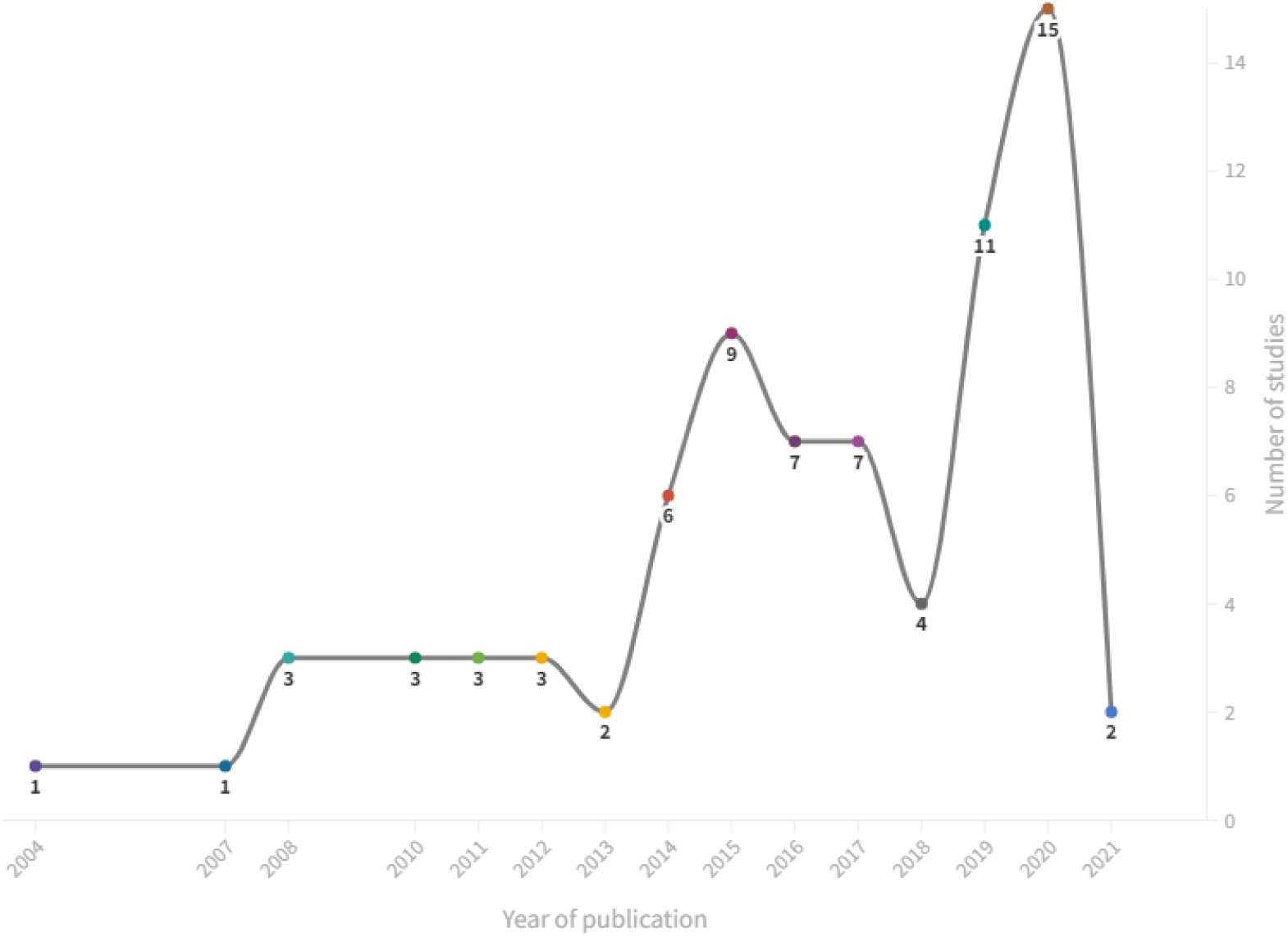
Publications per year of included studies (n=77). *The literature search was stopped in March 2021*. a combo graph (line plus column) plotting the number of studies included in the scoping review for each year. A progressive increase of publications occurred between 2004 and 2020.

#### Country of origin

The included publications were from 21 different countries. All continents were represented in this repartition. The most represented countries are USA (n = 11), Brazil (n = 9), Canada and India (n = 7). The complete geographical distribution of included studies is presented on the *figure 3*.

**Figure 3.**
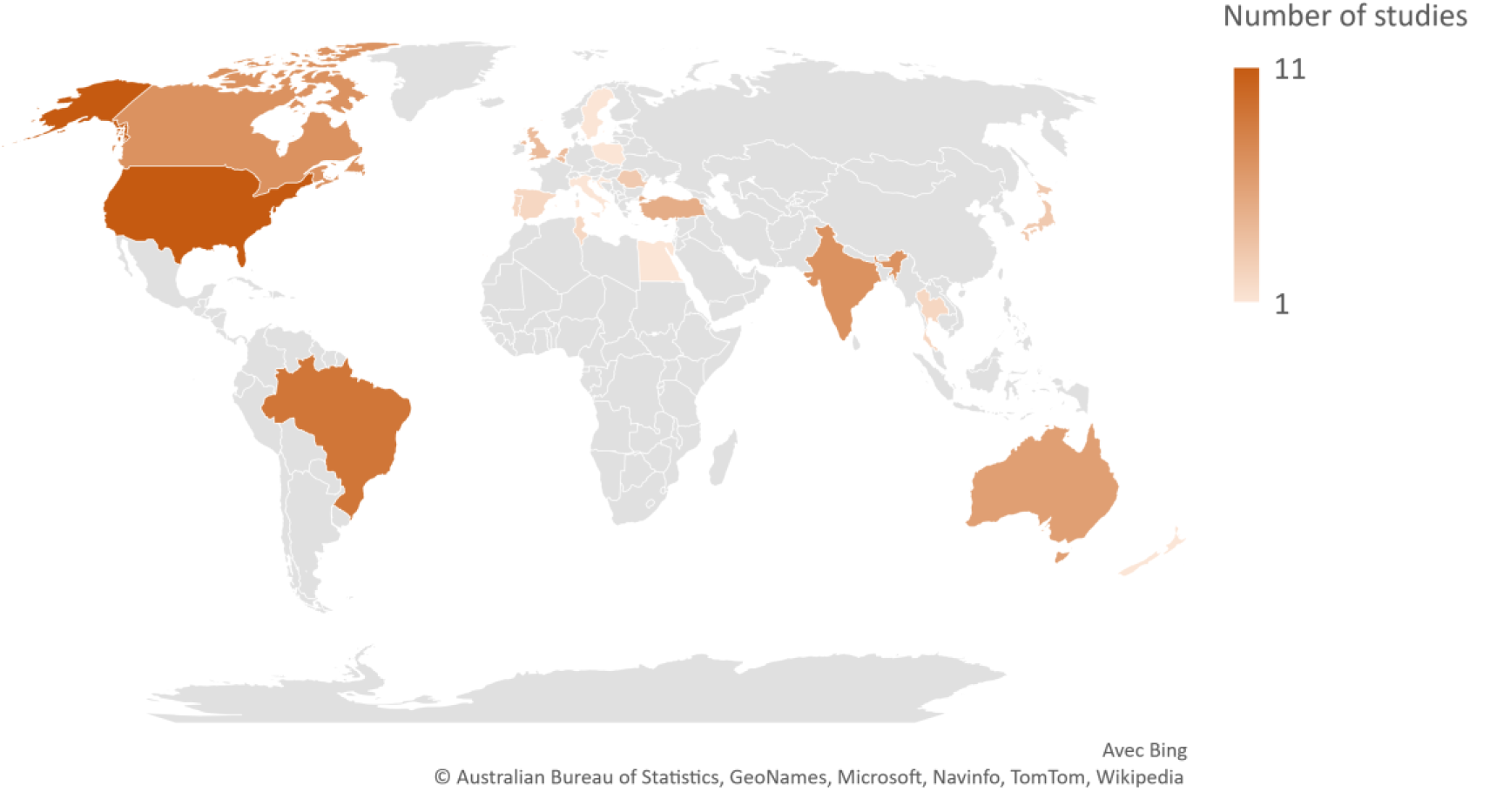
Worldwide geographical repartition of included studies (n = 77). a world map providing the geographical origins of studies included in the review. Countries with more studies were indicated by darker colour. USA, Brazil, Canada, Australia, India and Turkey were the most represented countries, respectively.

#### Article type

The majority of included studies were observational cross-sectional studies (n = 67). Other studies were cohort studies (n = 6), experimental pilot studies (n = 2), longitudinal study (n = 1) and reliability study (n = 1). Fifty-two studies included a control group to compare postural control of COPD patients to healthy subjects.

#### Participants

The number of participants ranged from 7 [24] to 1202 [25,26] with a median of 34. The median age for COPD patients was 68 years with a median forced expiratory volume in one second of 50 percent of the predicted value.

### Review findings

Overall evidence indicated that postural control is altered in COPD patients compared to healthy subjects. On the fifty-two studies that directly compare postural control of COPD patients with healthy subjects, forty-nine of them reported a statistically significant alteration in at least one postural control sub-component. This modification has been assessed by a wide variety of tools (both with laboratory and functional tools).

Every of the nine sub-components were assessed independently in the included studies. The complete distribution of the sub-components assessment in included studies is presented in the *figure 4*. The full detailed list of sub-components assessed in each included studies is available in the *appendix III*.

**Figure 4.**
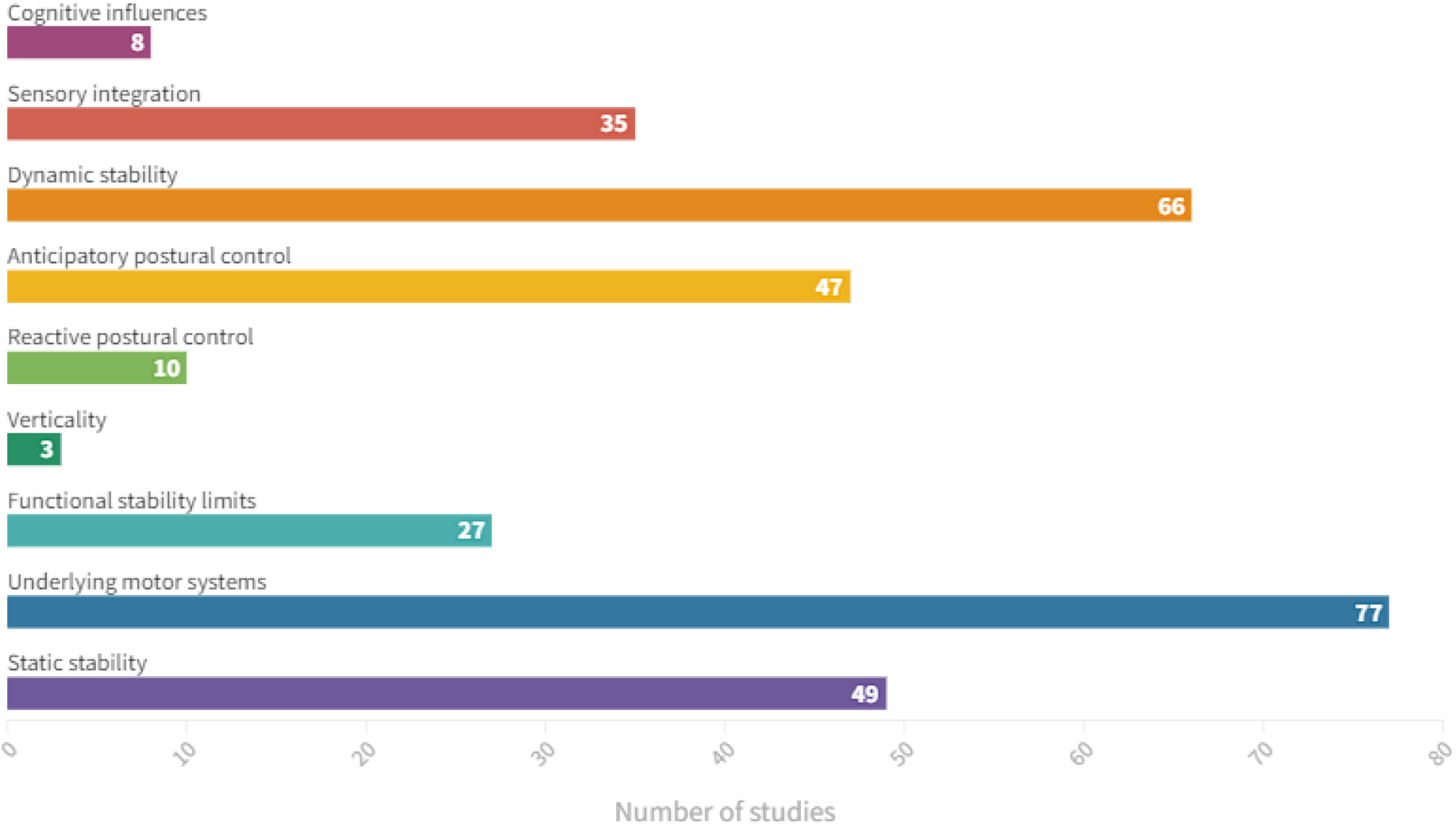
Repartition of postural control sub-components assessed in included studies (n=77). an horizontal bar chart plotting the number of studies that assessed each sub-component of postural control. The graph shows that all of the sub-components were assessed, with a minimum of three studies for “verticality” and a maximum of 77 studies for “underlying motor systems”. Long Description : an horizontal bar chart plotting the number of studies that assessed each sub-component of postural control. “Underlying motor systems” were assessed in 77 studies, then the following subcomponents were included in: 66 studies for “dynamic stability”, 49 studies for “static stability”, 47 studies for “anticipatory postural control”, 35 studies for “sensory integration”, 27 studies for “functional stability limits”, 10 studies for “reactive postural control”, 8 studies for “cognitive influences” and 3 studies for “verticality”.

#### Static stability

“Static stability” was assessed in forty-nine sources. The most frequent tools used to analyse “static stability” were force platform measurement, one leg stance test, specific parts of composite tests (BESTest, Berg Balance Scale (BBS), Tinetti, Short Physical Performance Battery (SPPB)) or the Sensory Organisation Test (SOT). In the included studies, “static stability” was assessed for several conditions of visual status (eyes open / eyes closed), stance (natural stance, tandem stance, one leg stance) or surface (stable / unstable surface, foam). A significant alteration of the component was found in the majority of studies: on thirty-five studies comparing COPD and healthy subjects, thirty studies reported a significant decrease in COPD group (twenty-six studies assessing “static stability” in isolation [26–51], four studies with composite tests [52–55]), four studies found no significant differences between groups [56–59] and data were not available in one study [60].

Nineteen studies used force platform measurements including an important variety of centre of pressure (COP) parameters [27–29,32,33,35,36,39–43,47,49,56,58,59,61,62]. Main COP variables used were area, velocity (in both anteroposterior (AP) and mediolateral (ML) directions), variability (AP and ML directions), and amplitude (AP and ML directions). Significant differences compared to healthy subjects were identified for each of these parameters, but results were heterogenous between studies [27–29,32,33,35,36,39–43,47,49]. The reported correlations between “static stability” and clinical factors are summarized in *table 2*.

**Table 2.**
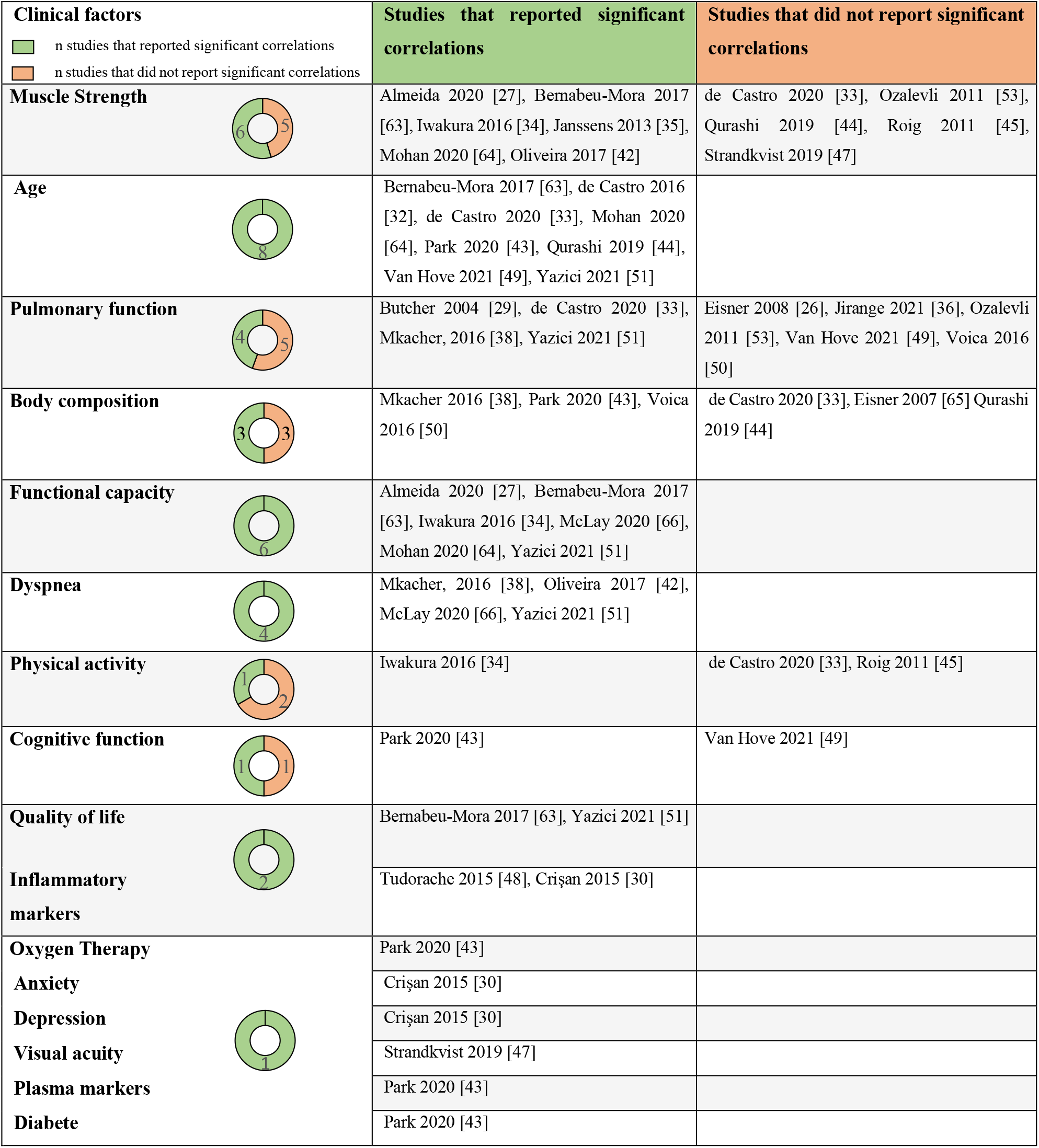
Summary of correlations between clinical factors and “static stability” sub-component.

#### Underlying motor systems

Using the operational definition of the “systems framework for postural control framework” [16], all the tools used in the included studies assess “underlying motor systems”. However, few studies offered the possibility to isolate the motor component of postural control. Electromyographic assessments were realized in 3 studies [33,54,62]. Beauchamp et al. [54] found no significant differences in lower limb muscles activation in COPD patients compared to healthy controls during a postural perturbation task, except for tibialis anterior onset. de Castro et al. [33] analysed the activation of neck, respiratory and hip muscles during different static conditions. They found a higher activation of scalene during one leg stance in COPD patients and a higher Gluteus Medius activation during eyes closed standing. Lastly, Smith et al. [62] reported that abdominal muscle activities (obliquus externus and rectus abdominis) were higher in COPD patients during postural tasks.

Some studies investigated the relative contribution of each postural control sub-component using the BESTest [44,54,55]. Studies by Beauchamp et al. [54] and Qurashi et al. [44] are consistent to identify the “biomechanics constraints” component of the BESTest (comprising “underlying motor systems”) as one of the most altered of those assessed, in COPD patients compared to controls. However, another study employing a similar design did not support that findings [55].

Studies by Liu et al.[67] and Janssens et al. [35] proposed stratified analyses of postural control, using low and high strength subgroups in COPD patients. Liu et al. [67] showed no differences in gait parameters between low and high quadriceps strength groups. On the other hand, Janssens et al. [35] demonstrated that COPD patients with low inspiratory muscle strength, have different postural control strategies compared to patients with higher inspiratory muscle strength, using a greater reliance on ankle-related proprioceptive signals and a more ankle-steered strategy.

#### Functional stability limits

Twenty-seven studies included an evaluation of “functional stability limits”. This sub-component was mainly assessed by the Functional Reach Test (FRT) (alone or included in a composite test). There is consistent and direct evidence of a significant decrease of limits of stability in patients with COPD compared to healthy controls [25,37,47,68]. One study showed that this alteration is predominant in the antero-posterior direction [47]. However, this finding was not supported by another study, that reported alterations in all directions tested [37]. Studies assessing postural control with composite functional tests (such as the BESTest or the BBS) also presented conflicting results: two studies [55,58] identified “functional stability limits” as one of the most impaired sub-component of postural control while other did not confirm these findings [44,54].

Concerning the associations with this sub-component, Eisner et al. [26] suggested that FRT was not associated with pulmonary function. Beauchamp et al. [54] found no association between this sub-component and different markers of lower limb muscle function or patient-reported physical activity. On the other hand, Standkvist et al. [47] reported that the major determinants of stability limits in the antero-posterior direction were muscle strength parameters (hip abductors, knee extensors, hand grip strength).

#### Verticality

“Verticality” was investigated in only three studies [39,54,55]. This component was assessed by sub-parts of composite balance tests (BESTest and mini BESTest). Among these sources, two studies allowed to isolate information on “verticality” [54,55], suggesting that this sub-component could be altered in COPD patients compared to healthy subjects. Relative comparison of the sub-components using the BESTest did not identify “verticality” as one of the most impaired postural control sub-components for COPD patients [54,55].

In the study by Beauchamp et al. [54], the “verticality / stability limits” component of the BESTest show no correlation with either daily-reported physical activity or lower limb strength.

#### Reactive postural control

Ten studies included assessments of “reactive postural control”. “Reactive postural control” was mainly assessed by stepping responses, in isolation or included in composite balance tests (eg.BESTest or BBS). Three studies found a specific impairment of this component in COPD patients compared to healthy subjects [44,54,55]. Comparative analysis of sub-components with the BESTest identified “reactive postural control” as one of the most impaired component of postural control in one study [55] but this was not supported by two other studies using a similar design [44,54].

Concerning the associations with the “reactive postural control” component, Beauchamp et al. [54] showed a significant moderate correlation with physical activity and a significant weak correlation with ankle strength.

#### Anticipatory postural control

“Anticipatory postural control” was assessed in forty-seven studies. This component was mainly analyzed by transitions assessment, in isolation or as a part of composite functional tests (e.g. BESTest or BBS) but also during simple functional tools such as the Timed up and Go test (TUG).

Only very few studies have the potential to give isolated information on this sub-component. Smith et al. [62] found no differences in the number of “anticipatory postural adjustments” between COPD group and controls, but a decreased ability for COPD patients to recover initial stability after the movement. In agreement with these findings, Beauchamp et al. [54] showed that if the frequency of anticipatory postural adjustments did not differ between COPD and controls, the duration of the anticipatory postural adjustments was longer in COPD patients. All studies using the BESTest as an outcome showed that the “anticipatory postural control” sub-component is significantly impaired in patients with COPD compared to healthy subjects [44,54,55]. One of these studies identified this sub-component as one of the most impaired among all [54], but it was not supported by the results of the two others [44,55]. Regarding correlations of this sub-component, Beauchamp et al. [54] found significant weak correlations with physical activity and knee flexion strength.

#### Dynamic stability

This component was assessed in sixty-six studies included in the review. “Dynamic stability” was evaluated by various tools: the Timed up and Go test (TUG), laboratory gait analysis and by composite balance tests (BESTest, BBS, SPPB) including a specific part for dynamic stability assessment.

All studies that used the TUG as an outcome reported a significant constant alteration compared to healthy subjects [30–33,36,39,45,48,50,51,68–74]. In addition, when a sub-component analysis is performed in studies using composite balance tests, the dynamic stability component was significantly impaired compared to control participants [44,54,55]. These three studies were consistent to identify dynamic stability sub-component as one of the most impaired among all postural control sub-component.

Some studies using laboratory gait analysis identified impairments in parameters related to “dynamic stability” in COPD patients: alteration of margins of stability (in medio-lateral direction) and its variability [75] and increase of centre of mass medio-lateral sway [60] and acceleration variability [76].

Moreover, eight studies reported significant differences in gait parameters when compared to healthy subjects [63,72–78]. It concerns spatio-temporal parameters (e.g. Gait speed, cadence or step time) [76–78,80,82], variability of spatio-temporal parameters (e.g. Coefficient of variation of step time or stride length) [67,77,82] and kinematics parameters (e.g. Ankle range of motion or power absorption at ankle level) [79,81].

Significant correlations were reported between dynamic stability and numerous clinical factors, and they are presented in the *table 3*.

**Table 3.**
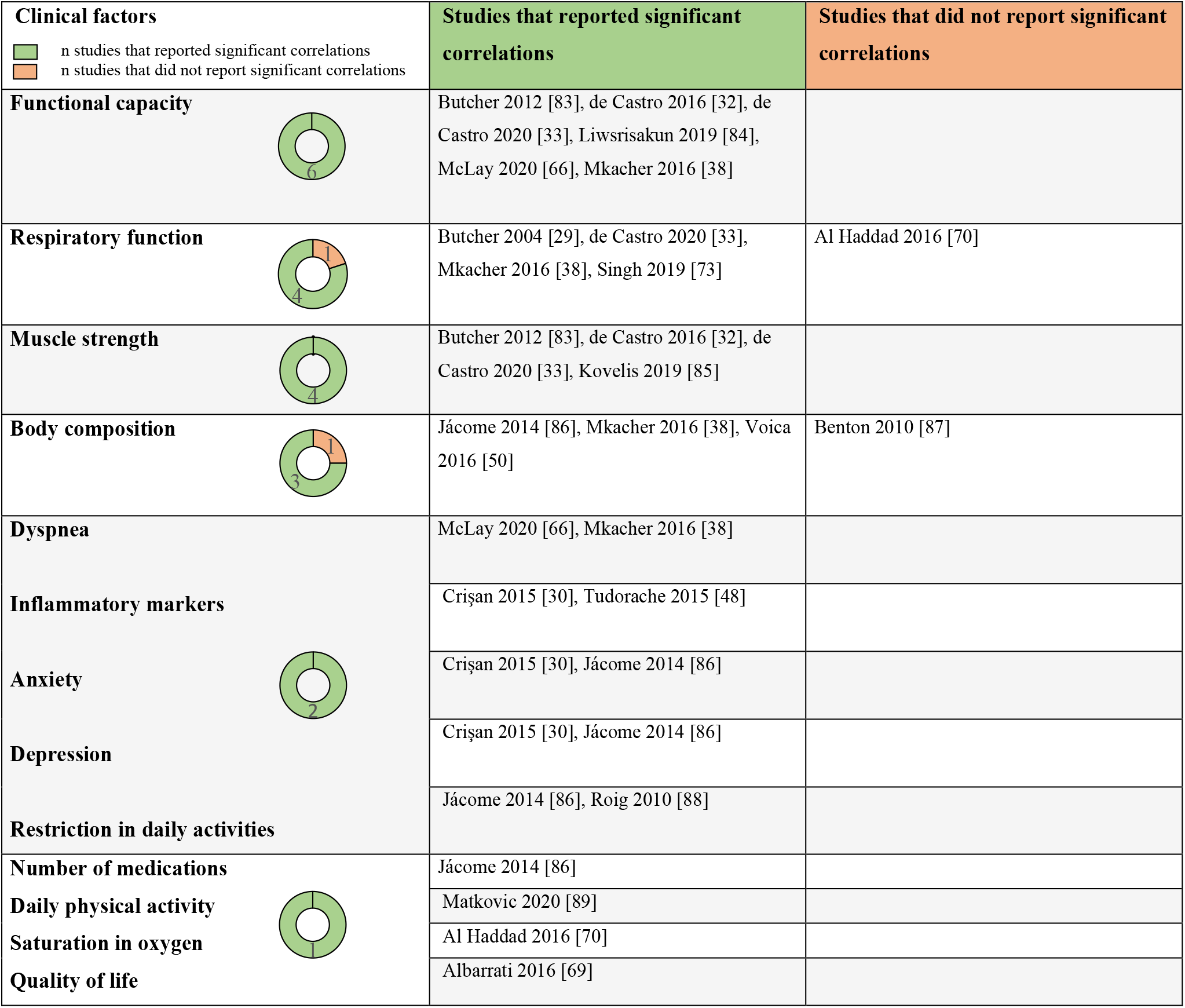
Summary of correlations between clinical factors and “dynamic stability” sub-component.

#### Sensory integration

“Sensory integration” was evaluated in thirty-five studies. “Sensory integration” was assessed by manipulating sensorial inputs (visual, vestibular, or somatosensory inputs) during different conditions (mainly in static condition, such as tandem stance or one leg stance). The SOT could be considered as a specific assessment tool of this sub-component and was employed in three studies [45,46,57]. This sub-component was also investigated in specific sub-parts of composite tools (eg.BESTest or BBS).

Studies that used the BESTest (or a derivative), were consistent to report that the sensory integration sub-component was impaired in COPD patients compared to healthy subjects [44,54,55]. Some studies used the SOT to isolate the relative “weight” of the different sensory inputs: Shalaby et al. [46] found that the overall SOT score is impaired in COPD patients compared to healthy controls, and that the conditions that targeted the vestibular system are altered. Roig et al. [45], using a similar design, also found a significant decrease in the total SOT score but did not identify significant differences in any of the conditions tested. In contrast, Pedrozo and Silveira [57] reported no significant difference between COPD group and control group using the SOT.

Some studies investigated the effect of the vision or the stability of the surface on postural control: Morlino et al. [39] reported no specific effect of the vision on postural control. Oliveira et al. [42] found no influence of the vision condition but that the surface (foam) caused a significant impairment in patients with COPD. Almeida et al. [27] tested various conditions (visual and surface) but reported no effect nor interaction between the conditions on the postural control. Strandkvist et al. [47] reported differences in unstable support condition eyes open whereas this difference was not found eyes closed. Butcher et al. [29] reported differences between COPD and healthy subjects only in the same condition (unstable surface, eyes open), no difference was found in other conditions.

In addition, Janssens et al. [35] found significant differences of proprioceptive pattern in COPD subjects compared to healthy controls : COPD patients increased their reliance on ankle muscle proprioceptive signals and decreased their reliance on back muscle signals.

Regarding the associations, Beauchamp et al. [54] reported that the sensory integration component of the BESTest was significantly correlated with lower limb strength (moderately with knee flexion strength and weakly with knee extension and ankle flexion).

#### Cognitive influences

“Cognitive influences” were assessed in eight studies. This sub-component was investigated using various cognitive task such as counting backwards or reciting a list of words during static or dynamic conditions (eg. on force plateform, during the TUG). Evidence on this component was sparse, and only few studies offered the opportunity to isolate information on cognitive influences and postural control of COPD patients. Cognitive tasks impair postural control performance in both COPD and healthy subjects [39,49,72]. Two studies [49,72] showed no supplementary effect of the cognitive task in COPD patients compared to controls while Morlino et al. [39] found the opposite. In addition, studies that assessed postural control by using the BESTest reported that the “cognitive influences” sub-component is impaired compared to healthy subjects [54,55]. Finally, the study by Van Hove et al. [49] reported that different types of cognitive tasks produced different effects on postural control in COPD patients.

Concerning the associations with this sub-component, one study found no association between cognitive ability and postural control assessed on Wii balance board [49]. Another study identified weak correlations between the double task-TUG and dyspnea and with self-reported physical functioning [66]. These correlations were similar with those reported for the TUG without additional cognitive task in the same study.

#### Postural control and activities of daily living

This review identified five sources that investigate the association between postural control of COPD patient and ADL. Cruz et al. [90] reported that COPD patients with a postural control impairment are more dependant in at least one activity of daily living (ADL) and are more severely restricted in daily life than patients without postural control impairment. Another study from the same team [86] identified determinants of “dynamic stability”: restriction in recreational activities was retained among other clinical factors (such as body mass index, depression score or number of medication) in a multiple regression model explaining around one third of the stability variance.

Three studies examined the correlations between postural control and various indicators related to ADL: Roig et al. [88] reported significant moderate correlations between “dynamic stability” (assessed by the TUG) and a specific ADL (stair climbing). In the study by Almeida et al. [27], a parameter of “static stability” was moderately correlated with the Glittre ADL (a specific functional assessment of ADL-related ability). Finally, Albarrati et al. [69] found a significant but weak correlation between “dynamic stability” (assessed by the TUG) and the activity score of the Saint-George Respiratory Questionnaire (a specific section focusing on difficulties during ADL).

## Discussion

This review examined the available evidence on postural control sub-components impairments and characteristics in patients with COPD. The associations between postural control and activities of daily living in this population were also explored. Seventy-seven studies were included indicating that this review is timely, permitting to identify gaps in knowledge and outlining interesting perspectives for research.

We will discuss the main results of the review for each sub-component and then the relation between postural control and activities of daily living.

### Static stability

Most studies underlined an impairment of static stability in people with COPD compared to healthy subjects. However, no specific pattern of alteration of “static stability” has been identified in the review. Indeed, in studies using force platform as an outcome, findings are various and heterogenous on centre of pressure parameters, preventing any firm conclusion. One potential explanation is that sub-groups analysis of COPD patients may be necessary to identify some homogenous phenotypes of static stability alterations. In addition, interesting associations between “static stability” and some clinical factors were reported, especially for dyspnea, age, and functional capacity where results of studies were consistent. If these associations are confirmed, they could help for sub-group analysis of static stability alterations. The relations between age or functional capacity and static stability have already been reported in general population [91,92]. Dyspnea is a cardinal symptom for COPD patients and its influence on postural control has been poorly studied. Dyspnea leads to an increase of neural drive, modifications of central nervous system regulation, changes of dynamic respiratory mechanics and has a psychological impact on patients [93]. All these alterations could potentially interfere with some sub-components implicated in postural control and lead to a worst performance for people with COPD.

### Underlying motor systems

Results of included studies support that underlying motor systems participate to postural control alteration in COPD patients. Moreover, some studies suggest that this sub-component is one of the most impacted compared to healthy subjects [44,54,55]. Muscle dysfunction is a well-established extra-respiratory impairment in COPD [94] and high-quality evidence showed that several muscle qualities are altered in this population [95]. As motor system is an important contributor to postural control [96] and muscle parameters are associated with postural control ability in healthy subjects [97], it could also play a central role in COPD postural control impairment [98]. In this review, included studies focused on muscular strength but other muscular qualities such as power and endurance could be relevant for postural control system [97,98].

This review showed that patients with COPD present different muscle activations patterns at both inspiratory, expiratory, and lower limb levels to maintain stability compared to healthy participants. These findings suggest that the precise assessment of these muscles could have an important role in postural control evaluation in COPD patients. Muscle function training is already a central feature of the treatment of COPD subjects [99] and could also play an interesting role in the management of postural control impairment.

### Functional stability limits

Available evidence indicates that this component is impaired in patients with COPD. In the elderly population, limits of stability are a valid predictor of future falls [100]. In COPD population, the alteration of limits of stability could be an important factor regarding the occurrence of falls. Conflicting evidence exists on the predominance of the direction(s) of the impairment and on the relative importance of this component alteration compared to others postural control components. Identification of direction-specific impairment could have some implication for the rehabilitation of patients [101]. Lower limb and hand grip strength seems to be major determinants of antero-posterior limits of stability in patients with COPD [47]. These findings suggest that the muscular strength is associated with functional stability limits and could be relevant for the management of COPD patients.

### Verticality

This sub-component of postural control is the less frequently assessed in the included studies. The few available results seem to indicate that verticality perception is impaired in COPD population. Previous research on the perception of verticality has highlighted the central role of brain areas (such as the thalamus) integrating inputs from visual, vestibular, and somatosensory systems [102,103]. In COPD patients, structural brain changes have been reported and can concern the thalamus [104]. These central modifications could be in relation to the impairment of verticality perception in COPD population. Based on relative comparisons reported in only two studies [54,55], verticality sub-component is not one of the most impacted by the disease. However, one study by Chauvin et al. [105] identified this sub-component as a robust predictor of falls in COPD patients in combination with the functional stability limits sub-component. This could underline the importance of integrating this sub-component in the assessment of postural control in COPD patients.

### Reactive postural control

This sub-component seems to be impaired in patients with COPD compared to healthy controls. In older adults, an impairment of reactive postural control represents an established risk factor of fall [106]. As the prevalence of falls have been reported to be high in the COPD population [42], factors influencing them are important to be identified.

Muscle function has an important influence on this sub-component: a systematic review conducted in older individuals, established that muscle fatigue induces deteriorations in reactive postural control [107]. Our review did not establish a direct link between muscle fatigue and this sub-component in COPD. However, an included study found a correlation between reactive postural control and lower limb strength [54] and muscle fatigue is a common and relevant disorder in patients with COPD [108].

### Anticipatory postural control

There is evidence in the literature showing that this sub-component can be impaired in people with COPD. Available evidence suggests that a longer duration of the anticipatory postural adjustments rather than the number of them could be the underlying mechanism of this impairment [54]. The duration of anticipatory postural adjustments is under the control of the cortical activity of central nervous system [109]. As structural brain changes have been documented in patients with COPD [104], it could have implications for “anticipatory postural control”. Fear of falling is also related to the impairment of this sub-component in other populations [110–112] and is often present in people with COPD [52].

### Dynamic stability

Dynamic stability was frequently studied in people with COPD. Results are consistent to report an alteration of “dynamic stability” in patients with COPD compared to healthy controls. This review also suggests that this sub-component could be one of the more impaired among postural control components [44,54,55]. The alteration of dynamic stability could be more important in the medio-lateral direction than in the antero-posterior direction [60,75,76]. As modifications in the medio-lateral direction are related to an increased risk of falls in elderly [113], this emphasizes the need of the assessment of this specific characteristic of “dynamic stability” in COPD patients.

Quantified motion analysis represents an interesting option for the purpose of an accurate understanding of postural control in COPD patients. Several studies employing this tool were included is the review, however they mainly focused on gait. Other relevant activities of daily living such as stairs climbing, transitions or objects carrying were not fully investigated. Investigating these tasks using motion analysis could give relevant information on the role of postural control in activities of daily living impairment in COPD population.

### Sensory integration

This review identify evidence showing that this sub-component is impaired in patients with COPD. However, sensory integration implicates different inputs and a central reweighting, and the precise changes related to each sensory input are still unclear. We found studies indicating that nature of the surface of support could induces more alterations of postural control than the manipulation of the visual input [42,47]. It could implicate that plantar sensory system and proprioceptive system are less performant in individuals with COPD and that the reliance to the visual input is not increase for these patients. Another possible explanation is that the reweighting of the sensory information is different in patient with COPD. This is in line with the study of Janssens et al. [35], indicating that the proprioceptive weighting of COPD patients is more related to lower limb information than trunk proprioceptive signals. In addition, the literature shown that COPD patients could have structural brain changes [104]. It could be hypothesised that these modifications could have an impact on the central integration and processing of sensory data.

### Cognitive influences

Various cognitive tasks induce changes in postural control of both healthy and COPD patients. However, there is conflicting evidence that cognitive tasks cause more alteration on postural control in COPD patients than in healthy subjects. Previous research with elderly people has shown an age-related decrease of postural control performance under dual task conditions [114]. It is currently unclear if COPD disease provokes an additional impairment, but some specific factors associated with the pathology could be of interest regarding cognitive influences and postural control: for example, dyspnea has a known negative impact on cognitive parameters [115]. The presence of both acute and chronic dyspnea could induce modifications on cognitive function with some potential consequences on postural control. Other prevalent clinical factors in COPD, such as pain, could produce relevant interaction with cognitive load and postural control [116].

### Postural control and ADL

Activities of daily living are often difficult for patients with COPD [117]. These difficulties have some consequences in patients’ quality of life [118]. Some studies suggest that a performant postural control system is associated with a good realization of activities of daily living [119,120]. COPD patients with postural control impairment could experiment more difficulties in the realization of these tasks [86,90]. Postural control impairment is not only relevant for its association with the risk of fall but could potentially lead to an increased metabolic cost of the movement [121]. As research has already established that COPD patients present an increased energy expenditure compared to healthy subjects [122], it is possible that postural control impairment contributes to it. Finally, it can be hypothesized that increasing the postural control capacities of COPD patients could lead to a better daily life tasks realization.

### Strength and limitations

To our knowledge this is the first review aiming to summarize evidence on sub-components of postural control in COPD population. Using a robust conceptual and clinically relevant model, this review is interesting both for researchers and clinicians. By incorporating grey literature, this work aims to propose a wide and realistic synthesis. The methodology of the review is in line with the JBI guidelines for scoping reviews that represents the actual reference for designing and carrying out this type of review.

As with any review, these results should be interpreted with the consideration of some limitations. The combination of interrogated databases was done to ensure a wide coverage of the literature; however, it is possible that relevant sources have been omitted as some databases combinations could be more performant. The literature search has been performed only in French and English, preventing the inclusion of sources in other languages. In addition, no critical appraisal was performed as it is not a requirement for scoping review.

### Perspectives and futures directions

From this review different research perspectives could be drawn. They are presented in the *table 4* for each postural control sub-component and for activities of daily living. If this review proposed perspectives for each sub-component, postural control is a complex system that use sub-components in perpetual coordination during real life. Therefore, the understanding of postural control impairment in COPD, its origins, and its consequences will require in the future a wide variety of both human (different professions and skills) and material resources (analysis tools from new technologies for example).

**Table 4.**
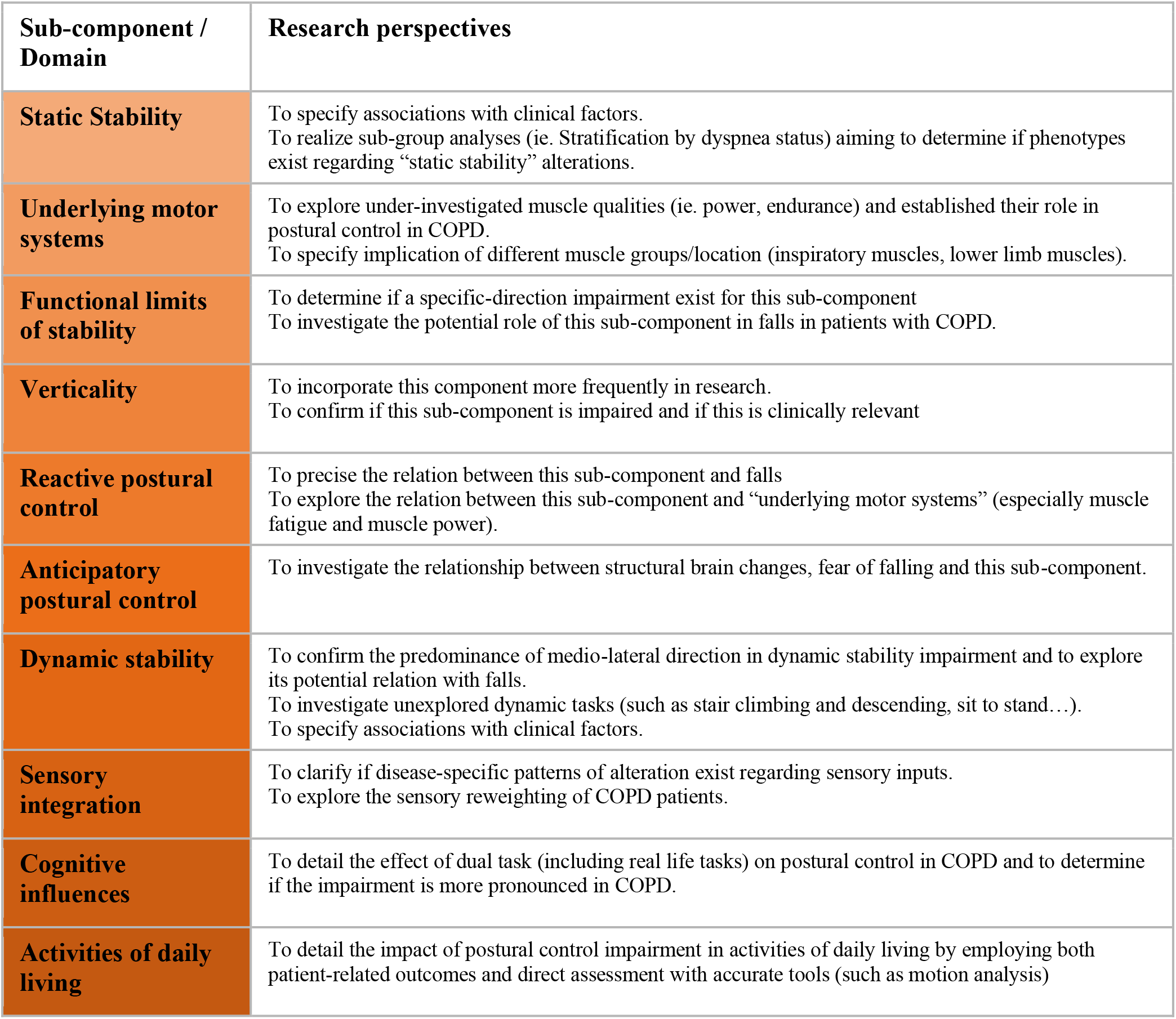
Research perspectives.

## Conclusion

This review provides an overview of the current knowledge on postural control sub-components in people with COPD. This work highlights that most of the postural control sub-components could be altered in the latter. Results underline the relative poverty of scientific literature concerning the relation between postural control of COPD patients and activities of daily living (perceived difficulties, performance, and characteristics of realization). New professional orientations and research perspectives will require comprehensive and multidisciplinary work including contributors from different fields to introduce new ways of postural control impairment assessment and management in COPD patients from an evidence-based perspective.

## Data Availability

All data produced in the present study are available upon reasonable request to the authors

## Acknowledgments

Olivier Honoré, librarian at IFPEK Rennes, for his support in literature search strategy development. Dr. Karim Jamal, researcher, and physiotherapist, for his expert input on literature review and postural control.

## APPENDICES

### Appendix I. Pubmed search strategy

| Search ID | Search terms | Results (n) |
| --- | --- | --- |
| #1 | ("postural balance"[MeSH Terms] OR "balance"[Text Word] OR "postural control"[Text Word]) AND (english[Filter] OR french[Filter]) | 248 118 |
| #2 | ("pulmonary disease, chronic obstructive"[MeSH Terms] OR "bronchitis, chronic"[MeSH Terms] OR "pulmonary emphysema"[MeSH Terms] OR "COPD"[Text Word] OR "chronic obstructive pulmonary disease"[Text Word]) AND (english[Filter] OR french[Filter]) | 76 717 |
| #3 | ("muscle, skeletal"[MeSH Terms] OR "muscle skeletal"[Text Word] OR "muscle strength"[MeSH Terms] OR "muscle strength"[Text Word] OR "paresis"[MeSH Terms] OR "muscle weakness"[MeSH Terms] OR "respiratory muscles"[MeSH Terms] OR "muscle activation"[Text Word]) AND (english[Filter] OR french[Filter]) | 297 371 |
| #4 | ("range of motion, articular"[MeSH Terms] OR "range of motion"[Text Word]) AND (english[Filter] OR french[Filter]) | 68 204 |
| #5 | ("cognitive dysfunction"[MeSH Terms] OR "cognitive dysfunction"[Text Word] OR "cognition"[MeSH Terms] OR "cognition"[Text Word] OR "executive function"[MeSH Terms] OR "executive function"[Text Word]) AND (english[Filter] OR french[Filter]) | 292 344 |
| #6 | ("gait"[MeSH Terms] OR "gait"[Text Word] OR "walking"[MeSH Terms] OR "walking"[Text Word]) AND (english[Filter] OR french[Filter]) | 120 674 |
| #7 | ("gravity sensing"[MeSH Terms] OR "orientation, spatial"[MeSH Terms] OR "postural orientation"[Text Word]) AND (english[Filter] OR french[Filter]) | 21 876 |
| #8 | ("proprioception"[MeSH Terms] OR "proprioception"[Text Word] OR "sensation"[MeSH Terms] OR "sensation"[Text Word] OR "sensation disorders"[MeSH Terms] OR "sensation disorders"[Text Word] OR "visual perception"[MeSH Terms] OR "visual perception"[Text Word] OR "visual acuity"[MeSH Terms] OR "visual acuity"[Text Word] OR "auditory perception"[MeSH Terms] OR "auditory perception"[Text Word] OR "hearing loss"[MeSH Terms] OR "hearing loss"[Text Word] OR "sensory reweighting"[Text Word]) AND (english[Filter] OR french[Filter]) | 702 550 |
| #9 | ("ankle strategy"[Text Word] OR "hip strategy"[Text Word] OR "multi joint coordination"[Text Word]) AND (english[Filter] OR french[Filter]) | 230 |
| #10 | ("activities of daily living"[MeSH Terms] OR "activities of daily living"[Text Word] OR "walking"[MeSH Terms] OR "walking"[Text Word] OR "lifting"[MeSH Terms] OR "lifting"[Text Word]) AND (english[Filter] OR french[Filter]) | 216 584 |

| Search ID | Query | Results (n) | Date |
| --- | --- | --- | --- |
| S1 | #1 AND #2 | 793 | 02/03/2021 |
| S2 | #2 AND #3 | 2423 | 02/03/2021 |
| S3 | #2 AND #4 | 22 | 02/03/2021 |
| S4 | #2 AND #5 | 461 | 02/03/2021 |
| S5 | #2 AND #6 | 1932 | 02/03/2021 |
| S6 | #2 AND #7 | 66 | 02/03/2021 |
| S7 | #2 AND #8 | 539 | 02/03/2021 |
| S8 | #2 AND #9 | 0 | 02/03/2021 |
| S9 | #1 AND #2 AND #10 | 64 | 02/03/2021 |
| S10 | #2 AND #10 | 3477 | 02/03/2021 |

### Appendix II. Charting table

| Charting table |  |
| --- | --- |
| Scoping review details |  |
| Scoping review title | Postural control and its sub-components in patients with copd : a scoping review protocol |
| Scoping review objectives | identify and characterize alterations of sub-components that impact postural control in patients with COPD.<br>The second aim of this scoping review is to clarify the association between postural control and activities of daily living in patients with COPD. |
| Scoping review questions | 1. Which sub-components of postural control are implicated in its alteration in COPD patients ?<br>2. What are the characteristics of these sub-components ?<br>3. What is the relation between postural control (globally and for each sub-components) and ADL in COPD patients ? |
| Eligibility criteria |  |
| Population | This review considered the studies that include COPD patients as defined by the Global Initiative for Obstructive Lung Disease (GOLD). All stages of the disease were considered. Patients must be over 18 years old. This review excluded studies involving other chronic respiratory lung diseases (eg. Asthma, interstitial lung diseases or bronchiectasis). |
| Concept | The concept of postural control (Systems Framework for Postural Control) proposed by Horak and his sub-components were explored in this review<br>This model was used and adapted by Sibley et al. who established an operational definition of the model. The adaptation includes 9 sub-components which are presented and defined in the table1.: 1. Static stability, 2. Underlying motor systems, 3. Functional stability limits, 4. Verticality, 5. Reactive postural control, 6. Anticipatory postural control, 7. Dynamic stability, 8. Sensory integration, 9. Cognitive influences.<br>This review included studies that focus on postural control, or at least one of its sub-components but in relation with global postural control strategies or performance in static or dynamic conditions. We considered as postural control assessment all functional tools described by Sibley et al in their review and laboratory assessment (kinetic or kinematic analyses in static and dynamic conditions). Subjective assessments of postural were not included in the review. Sources analysing one sub-component in isolation, with no direct link to postural control are excluded to the review. |
| Context | This review focused on available evidence on postural control with no geographical restriction. We included studies in the widest possible range of settings (laboratory assessment, inpatient or outpatient studies, pulmonary rehabilitation settings) and population (community-dwelling, real life...). |
| Types of sources | **observational studies** (descriptive studies, cohort studies, cross-sectional studies, case-control studies)<br>**experimental studies** (randomized controlled trials, non-randomized controlled trials and quasi-experimental designs such as before-after studies)<br>**systematic reviews.**<br>**Conference abstracts** and **grey literature** will be considered for inclusion |
| Evidence source details and characteristics |  |
| Citation details: |  |
| Title |  |
| Authors |  |
| Journal and Year |  |
| DOI / URL |  |
| Country |  |
| Context |  |
| COPD Participants<br>(number / age / %FEV1) |  |
| Controls (number / age) |  |
| Study methods |  |
| **Details / results extracted from source of evidence** |  |
| Postural control and sub-components analysed in the study (as defined in the protocol)<br>*bold if assessed in the study* | 1. Static stability |
|  | 2. Underlying motor systems |
|  | 3. Functional limits of stability |
|  | 4. Verticality |
|  | 5. Reactive postural control |
|  | 6. Anticipatory postural control |
|  | 7. Dynamic stability |
|  | 8. Sensorial integration |
|  | 9. Cognitive influences |
| Outcomes (and assessment methods) |  |
| Sub-components conclusions |  |

### Appendix III. Complete list of included studies with sub-components assessed in each study

| Studies | 1 Static stability | 2 underlying motor systems | 3 functional stability limits | 4 verticality | 5 reactive postural control | 6 anticipatory postural control | 7 dynamic stability | 8 sensory integration | 9 cognitive influences |
| --- | --- | --- | --- | --- | --- | --- | --- | --- | --- |
| Jirange 2021 | Yes | Yes | no | no | no | yes | yes | yes | no |
| Yazici 2020 | Yes | Yes | no | no | no | yes | yes | yes | no |
| Van Hove 2021 | Yes | Yes | no | no | no | no | no | yes | yes |
| Araujo de Castro 2020 | Yes | Yes | yes | no | yes | yes | yes | yes | no |
| Ozsoy 2020 | No | Yes | no | no | no | yes | yes | no | yes |
| Liwsrisakun 2020 | Yes | Yes | yes | no | no | yes | yes | yes | no |
| McLay 2020 | Yes | Yes | yes | no | yes | yes | yes | yes | yes |
| Park 2020 | Yes | Yes | yes | no | no | yes | yes | yes | no |
| Mohan 2020 | Yes | Yes | no | no | no | no | yes | no | no |
| Fallatafthi 2020 | No | Yes | no | no | no | no | yes | no | no |
| Singh 2019 | No | Yes | no | no | no | yes | yes | yes | yes |
| Bofino 2019 | Yes | Yes | no | no | no | no | no | yes | no |
| Liwsrisakun 2019 | Yes | Yes | yes | no | no | yes | yes | yes | no |
| Vardar-Yagli 2019 | No | Yes | no | no | no | yes | yes | no | no |
| Terui 2018 | Yes | Yes | no | no | no | yes | yes | no | no |
| Leung 2019 | Yes | Yes | yes | no | yes | yes | yes | yes | no |
| Bernabeu-Mora 2017 | Yes | Yes | no | no | no | no | yes | no | no |
| Morlino 2017 | Yes | Yes | no | yes | yes | yes | yes | yes | yes |
| Oliveira 2017 | Yes | Yes | yes | no | no | yes | yes | yes | no |
| **Araujo de Castro 2016** | Yes | Yes | no | no | no | yes | yes | no | no |
| **Iwakura 2016** | Yes | Yes | no | no | no | no | yes | no | no |
| **Voica 2016** | Yes | Yes | yes | no | no | yes | yes | yes | no |
| **Smith 2016** | No | Yes | no | no | no | yes | no | no | no |
| **Tudorache 2015** | Yes | Yes | yes | no | no | yes | yes | yes | no |
| **Yumrutepe 2015** | No | Yes | yes | no | no | yes | yes | no | no |
| **Nantsupawat 2015** | No | Yes | no | no | no | no | yes | no | no |
| **Crisan 2015** | Yes | Yes | yes | no | no | yes | yes | yes | no |
| **Oliveira 2015** | Yes | Yes | yes | no | no | yes | yes | yes | no |
| **Cruz 2015** | No | Yes | no | no | no | yes | yes | no | no |
| **Janssens 2014** | No | Yes | no | no | no | yes | yes | no | no |
| **Jacome 2014** | No | Yes | no | no | no | yes | yes | no | no |
| **Janssens 2013** | Yes | Yes | no | no | no | no | no | yes | no |
| **de Buyser 2013** | Yes | Yes | no | no | no | yes | yes | no | no |
| **Roig 2011** | Yes | Yes | no | no | no | no | no | yes | no |
| **Ozalevli 2011** | Yes | Yes | yes | no | no | yes | yes | yes | no |
| **Beauchamp 2012** | Yes | Yes | yes | yes | yes | yes | yes | yes | yes |
| **Rocco 2011** | Yes | Yes | yes | no | yes | yes | yes | yes | no |
| **Smith 2010** | Yes | Yes | no | no | no | no | no | no | no |
| **Eisner 2008** | Yes | Yes | yes | no | no | no | yes | no | no |
| **Chang 2008** | Yes | Yes | no | no | no | yes | yes | no | no |
| Eisner 2008b | Yes | Yes | yes | no | no | no | yes | no | no |
| Eisner 2007 | Yes | Yes | no | no | no | no | yes | no | no |
| Butcher 2004 | Yes | Yes | yes | no | no | yes | yes | yes | yes |
| Porto 2017 | Yes | Yes | yes | no | no | yes | yes | yes | no |
| Butcher 2012 | No | Yes | no | no | no | yes | yes | no | no |
| Roig 2010 | No | Yes | no | no | no | yes | yes | no | no |
| Benton 2010 | No | Yes | no | no | no | yes | yes | no | no |
| Liu 2020 | No | Yes | no | no | no | no | yes | no | no |
| McCamley 2017 | No | Yes | no | no | no | no | yes | no | no |
| Matkovic 2020 | No | Yes | no | no | no | yes | yes | no | no |
| Al Haddad 2016 | No | Yes | no | no | no | yes | yes | no | no |
| Yentes 2015 | No | Yes | no | no | no | no | yes | no | no |
| Yentes 2014 | No | Yes | no | no | no | no | yes | no | no |
| Annegarn 2012 | No | Yes | no | no | no | no | yes | no | no |
| Pedrozo 2015 | Yes | Yes | no | no | no | no | yes | yes | no |
| Mkacher 2016 | Yes | Yes | yes | no | yes | yes | yes | yes | no |
| Lee 2018 | Yes | Yes | no | no | no | no | no | no | no |
| Mkacher 2014 | Yes | Yes | yes | no | yes | yes | yes | yes | no |
| Almeida 2020 | Yes | Yes | no | no | no | no | no | yes | no |
| Champion 2019 | Yes | Yes | yes |  |  | yes | yes | yes | no |
| Sayed 2021 | Yes | Yes | no | no | no | yes | yes | yes | no |
| Hetal 2018 | Yes | Yes | yes | yes | yes | yes | yes | yes | yes |
| **Kamble 2020** | Yes | Yes | yes | no | no | yes | no | yes | no |
| **Strandkvist 2019** | Yes | Yes | yes | no | no | no | no | yes | no |
| **Shalaby 2019** | Yes | Yes | no | no | no | no | no | yes | no |
| **Carter 2020** | No | Yes | yes | no | no | yes | yes | no | no |
| **Albarrati 2016** | No | Yes | no | no | no | yes | yes | no | no |
| **Qurashi 2019** | Yes | Yes | yes | no | yes | yes | yes | yes | no |
| **Liu 2017** | No | Yes | no | no | no | no | yes | no | no |
| **Yentes 2017** | No | Yes | no | no | no | no | yes | no | no |
| **Iwakura 2019** | No | Yes | no | no | no | no | yes | no | no |
| **Kovelis 2019** | No | Yes | no | no | no | yes | yes | no | no |
| **Medina-Mirapeix 2018** | Yes | yes | no | no | no | no | yes | no | no |
| **Patel 2014** | yes | yes | no | no | no | no | yes | no | no |
| **Lage 2020** | Yes | Yes | no | no | no | no | yes | no | no |
| **Lahousse 2015** | No | Yes | no | no | no | yes | yes | no | no |
| **Rutkowski 2014** | No | Yes | no | no | no | no | yes | no | no |

